# A simple mathematical model and physical device to estimate a woman-specific probability of skilled birth assistance and associated benefit of maternity waiting home stay

**DOI:** 10.1101/2024.04.02.24305221

**Authors:** Jérémie Gallien, George Chen, Yi Zhang, Yuhang Du, Jody Lori, Joseph Sieka, Bentoe Tehoungue

## Abstract

This paper presents a simple mathematical model and an associated physical device to predict (i) the risk that a woman’s active labor will begin without a skilled birth attendant based on her parity and anticipated time to access skilled care; and (ii) the extent to which that risk may be reduced by moving to a maternity waiting home some time before her expected due date. This tool is designed to facilitate more systematic discussions and better-informed decisions about labour care access arrangements during antenatal consultations.

## 1. Introduction

Motivated by the high maternal mortality ratio in many low-income regions of the world (e.g., 531 per 100,000 live births in Sub-Saharan Africa in 2020), Sustainable Development Goal 3.1 calls for the reduction of that ratio to 70 per 100,000 live births globally by 2030^2^. Among multiple interventions known to be effective against maternal mortality, the presence of a skilled birth attendant (SBA) is recognized to be material^1,3,4^ and thus tracked as Indicator 3.1.2 along with maternal mortality ratio (Indicator 3.1.1) as part of the Sustainable Development Goals initiative^2^.

The ease and speed of access to an SBA varies substantially across women based on their living location and access to transportation, local road infrastructure and seasonal conditions, the location of nearby health facilities and trained personnel and other factors^1,5^. The duration of the latent phase of labour from onset until the beginning of the active phase, when an SBA becomes more critical and transportation more challenging, also varies substantially across women^6-12^. For women at risk of delivering without an SBA, Maternity Waiting Homes (MWHs) situated near healthcare facilities and/or staffed by trained personnel may constitute a beneficial care pathway, and are rapidly developing^13-18^. Some barriers to MWH use include food security, insufficient capacity, home responsibilities and decision autonomy however.

The decisions affecting labour care pathway that are faced by pregnant women in resource-limited settings (for example planned birth location, transportation arrangements, care provider engagement, planned move to health facility including MWH, arrangements for family work including income generation, food & water provision and child care) are thus potentially material to health, cognitively complex and psychologically difficult^5, 19-21, 38^; these decisions may also be strongly influenced by cultural beliefs and traditions. In this context we observe that pregnant women and those advising or caring for them typically have limited or no access to systematic guidance and information relevant to those decisions, let alone quantitative and/or evidence-based information. To address this gap we have developed a simple mathematical model that can be embodied by an inexpensive physical device and provides a quantitative estimate of a pregnant women’s risk of experiencing labour without an SBA based on some of her individual circumstances. More specifically, our model’s estimate is driven by her predicted time to access an SBA once labour starts, her parity and her decision of when to move to an MWH if applicable. It is envisioned that this tool may facilitate more systematic discussions and better-informed decisions concerning labour care access arrangements when used by pregnant women and care professionals as part of the antenatal care process.

The remainder of this document contains a description of this mathematical model (§2), its input data sources and output tables (§3), our proposed implementing physical device (§4) and a discussion of the limitations and opportunities associated with this work (§5).

## 2. Mathematical Model

The model to be described next provides a quantitative estimate of the probability that an individual pregnant woman will be assisted by an SBA when the active phase of her labour begins, which we denote ***P***(SBA) It considers the following input:

- ***t*:** estimated time to access an SBA from the first signs of labour onset;
- ***K*** ∈{*multiparous,multiparou*}: pregnant women parity;
- ***G***_*k*_(.): cumulative probability distribution function of the latent labour phase duration for women with parity ***k***. That is, ***G*** _***k***_ **(*x*)** is the probability that the duration of her latent phase of labour will be shorter than any given number ***x*** ≥0;
- ***d***: time when a pregnant woman plans to move to an MWH in relation to her expected due date (EDD), if applicable. For example, ***d*** = - 7 days if a woman plans to move to an MWH 7 days before her EDD. If a woman does not plan to stay in a MWH at all, we set ***d*** = +∞ by convention;
- ***F***(.): cumulative probability distribution function of the difference between actual delivery date (ADD) and estimated delivery date (EDD). That is, ***F(x)*** is the probability that a woman’s labour onset will begin before day EDD + ***x*** for any given number ***x*** representing a number of days (positive or negative) relative to the EDD, and we set ***F***(+∞) = 1 by convention.

Our mathematical model is defined by the following equation:

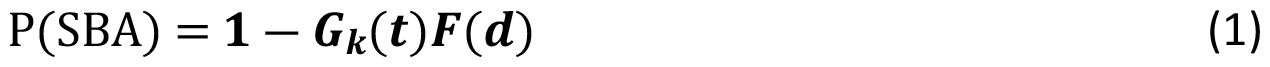

Note that, by the convention stated above, this equation simplifies to ***p***(SBA) =**1 ― *G***_***k***_ **(*t*)** for a woman not using an MWH. In this first simpler case, the equation thus expresses that the probability of an SBA not being present when the active phase of labour starts corresponds to the event that the duration of the latent phase of labour experienced by that woman is shorter than ***t***. In the second case when a woman moves to an MWH on day EDD + ***d***, equation (1) above expresses that the failure of an SBA to be present when the active phase of labour starts must result from the combined occurrence of the two following events: (i) the woman’s labour onset occurs before time EDD + ***d***, which has probability ***F*(*d*)** (i.e., her labour starts before her planned move to the MWH); and (ii) the woman’s latent phase duration is shorter than ***t***, which has probability ***Gk***(***t***) (i.e., after her labour starts she does not reach the nearest local SBA before the beginning of her active labour). Note that our model assumes that the onset of labour and the latent phase duration are independent, see §5 for a discussion of this and other model assumptions. Equation (1) thus captures and quantifies a key feature of the public health benefit of MWHs, namely that the initial risk of delivering without an SBA that is specific to a woman’s location and parity (as captured by the term ***G*** _k_ (***t***), which is larger for multiparous women and women with longer travel distance ***t***) can be mitigated in a commensurate manner by the time before her EDD when she moves to an MWH (as captured by the term ***F***(***d***), which will decrease towards zero with an earlier MWH move date EDD + ***d***, potentially reducing the overall risk term G _k_ (***t***) ***F***(***d***) arbitrarily close to zero). This is intuitive and consistent with our field observations that some MWHs will advise women in remote locations to come to the MWH earlier in their pregnancy than others living closer to a health facility.

## 3. Input Data Sources and Output Tables

In this section we first discuss our selection of the two main generic data input ***F***(.) and ***G*** _***k***_ (.) required by our model based on the existing clinical literature (§3.1 and §3.2), then provide tables of corresponding output data ***P***(SBA) predicted by our model (§3.3).

### 3.1 Labour Onset Distribution *F*(.)

Gestational age is the term used to describe the age of the foetus or the duration of the pregnancy. Main current methods for estimating human gestational age and expected remaining pregnancy duration or EDD are based on either ultrasound scans or the date of the last menstrual period (LMP), the latter being more common in resource-limited settings^22-24^. With either prediction method however, substantial variability between ADD and EDD remains. This is due to factors including inaccurate recollection of the LMP date, amenorrhea, variations in individual menstrual cycle lengths, use of oral contraceptive pills, irregular ovulation patterns, and the biological variability of foetal development^25-27^. We identified several studies reporting empirical findings on the difference between the ADD and LMP-based EDD predictions. Khambalia et al. assessed this variation by examining 10,243 women who underwent spontaneous labour and gave birth to a single child without significant anomalies^28^. Among other results Khambalia et al. found that the difference ADD – EDD had a mean of -1.48 days and a standard deviation (SD) of 9.21 days, that the ADD coincided with the EDD in about 5% of cases, and that approximately 66% of deliveries occurred within a one-week margin (±7 days) around EDD^28^. This is consistent with Tunon et al., who found that 56% of deliveries occurred within a one-week margin (±7 days) in a cohort of 9240 women with reliable LMP and spontaneous birth^29^. From a dataset of 34,249 singleton pregnancies from East Midlands Obstetric Database, Mongelli et al. found that the difference between ADD and EDD had a mean of -1.79 days with an SD of 11.3 days^30^. Failing to find an empirical study of ADD – EDD in a low-income setting close to that motivating our work however, we decided for now to use the data from Khambalia et al.^28^ in order to estimate our model input F(.). This is because that particular study reported a complete estimated empirical distribution (as opposed to summary statistics) for ADD – EDD, and relied on precise ADD data. Based on the empirical data from Khambalia et al.^28^, we estimated a continuous distribution for actual labour onset relative to EDD using Gaussian kernel density estimation^31,32^ with a bandwidth parameter of 0.3 and value range from -30 days to +30 days. The density and cumulative distribution functions corresponding to both the original empirical data and our estimated continuous distribution are shown in Figure 1.

**Figure 1:**
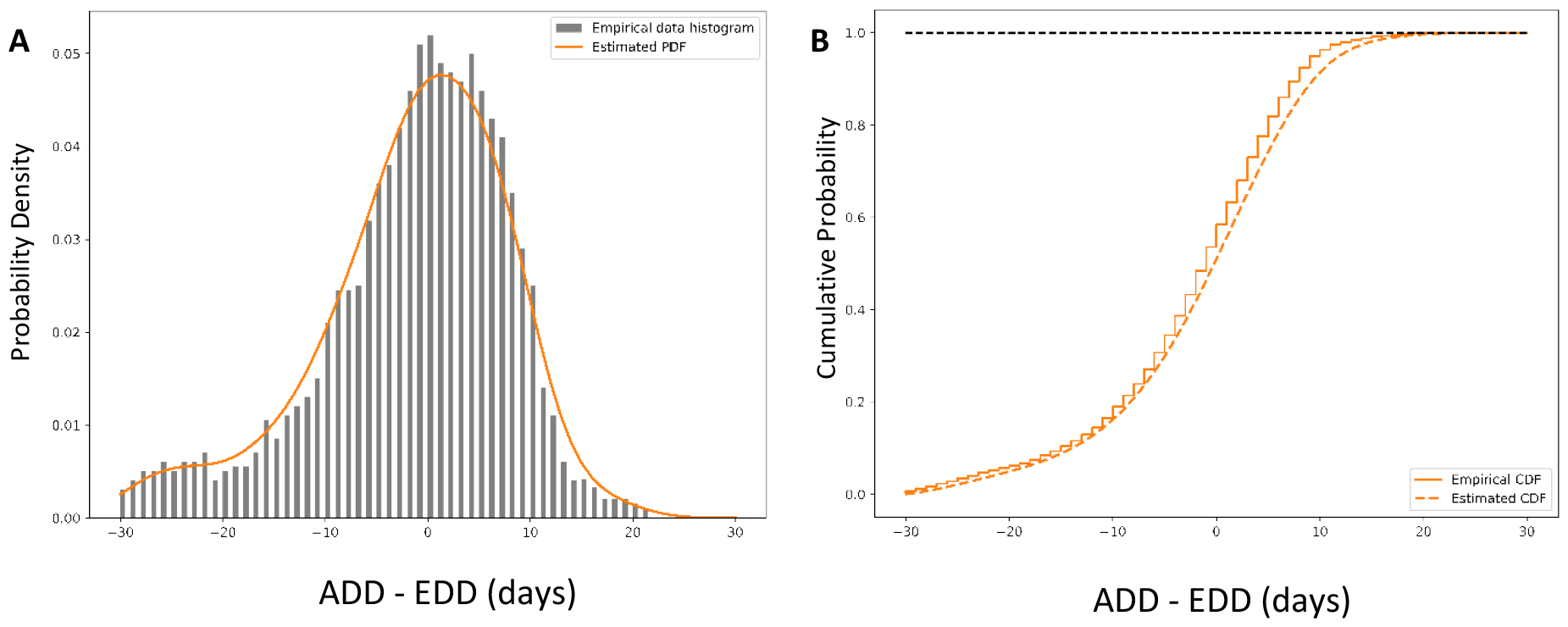
Empirical distribution and estimated continuous distribution of the difference between Actual Due Date (ADD) and Estimated Due Date (EDD) determined from the Last Menstrual Period (LMP) method, as estimated from Khambalia et al. ^28^ Panel A: Probability density Panel B: Cumulative probability under the empirical distribution and estimated continuous distribution (providing input data ***F***(.) in our model).

### 3.2 Latent Phase Duration Distribution G _*k*_ (.)

The latent phase is the interval between the onset of labour and the beginning of the active phase. The latent phase is clinically characterized by the presence of contractions, cervical dilation less than 6 cm upon examination and slow cervical dilation change. A definitive consensus on the precise definition of both beginning and end of the latent phase remains elusive, though it is widely held that a dilation of the cervix at or beyond 6 cm with concurrent contractions typically signifies a transition to the active phase of labor^6,7^. A seminal study of the human phases of labour was performed by Friedman, who defined the onset of the latent phase as the time when the patient felt significant, regular uterine contractions and a slow progression in cervical dilation noted by a clinician, and the end of the latent phase as the time of noticeable acceleration of cervical dilation over time^8,9^. Friedman reported complete empirical distribution of the latent phase estimated from these definitions, and in particular estimated the 95^th^ percentiles for the duration of the latent phase of spontaneous labour as 20 hours in nulliparas and 14 hours in multiparas^8,9^. More recent studies suggest that many women experiencing a latent phase exceeding the traditionally accepted normal duration stemming from Friedman’s work can still have a normal active phase and vaginal birth^33^; a recent prospective study including nearly 1300 healthy participants noted that the latent phase was approximately 10 hours longer than Friedman had previously noted, suggesting that patients may benefit from updated and relaxed expectations for what constitutes “normal” labour not requiring heavy clinical interventions^34^. We note that the exact clinical definition of the latent phase varies across studies^35^. Furthermore, these different definitions may still differ from the reality that our model construct ***G*** _***k***_ (.) seeks to represent, namely the time afforded to a woman from the decision to seek care after realising that her labour has started until the time when it becomes difficult for her to move and/or the presence of an SBA is material. For these reasons we conservatively decided for now to estimate ***G*** _***k***_ (.) from the published data suggesting the shortest latent phase durations, namely that reported by Friedman (mean 7.1 hours and SD of 4.0 hours for nulliparas, mean 5.3 hours and SD of 4.1 hours for multiparas)^8,9^.

We also employed the Gaussian kernel density method^,32^ to estimate continuous latent phase duration distributions for our model, using a bandwidth parameter of 0.3 and value ranges set to 0–30 hours for nulliparous women and 0–16 hours for multiparous women. The corresponding density and cumulative distribution functions associated with both the original empirical data and the estimated continuous distributions are shown in Figure 2.

**Figure 2:**
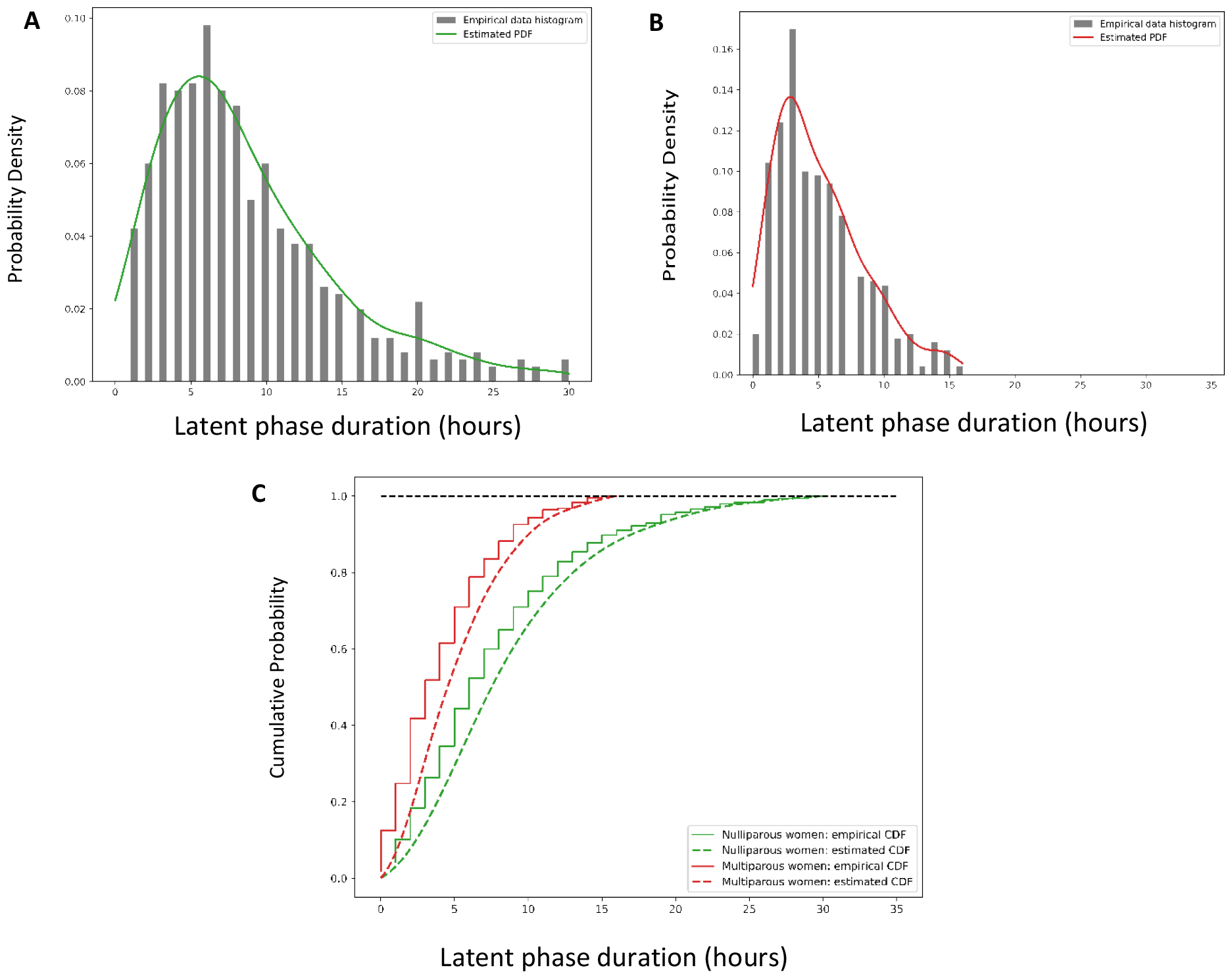
Empirical distributions and estimated distributions of the latent phase duration for nulliparas and multiparas, estimated from the data reported in Friedman. ^8,9^ Panel A: Probability density for nulliparous women Panel B: Probability density for multiparous women Panel C: Cumulative probability for both nulliparous and multiparous women. The estimated continuous cumulative probability functions provide input data ***G*** _***k***_ (.) in our model.

### 3.3 Output Data Tables

Tables 1 (nulliparas) and 2 (multiparas) show the output of the mathematical model defined in §2 when applied to the selected input data ***F***(.) and ***G*** _k_ (.) described in §3.1 and §3.2, respectively; these tables are constructed to illustrate how this mathematical model may be used to inform care pathway decisions for individual pregnant women and indeed contain the underlying data of our proposed physical device to be described in §4. Because the estimation by pregnant women of their travel time ***t*** from their home to the nearest health facility may be associated with some uncertainty, we consider different intervals of length equal to either 30 minutes or 1 hour for these travel times. Furthermore, we conservatively report for each travel time interval the lowest value of ***P***(SBA) within the interval, which is associated with the highest travel time (upper bound) that it contains. Finally, the MWH use scenarios we consider include ***d*** = +∞ (no MWH stay) and ***d*** ∈ {0, ― 7, ― 14, ― 21, ― 28} (move to an MWH on the EDD and 1, 2, 3 and 4 weeks before the EDD, respectively).

**Table 1:**
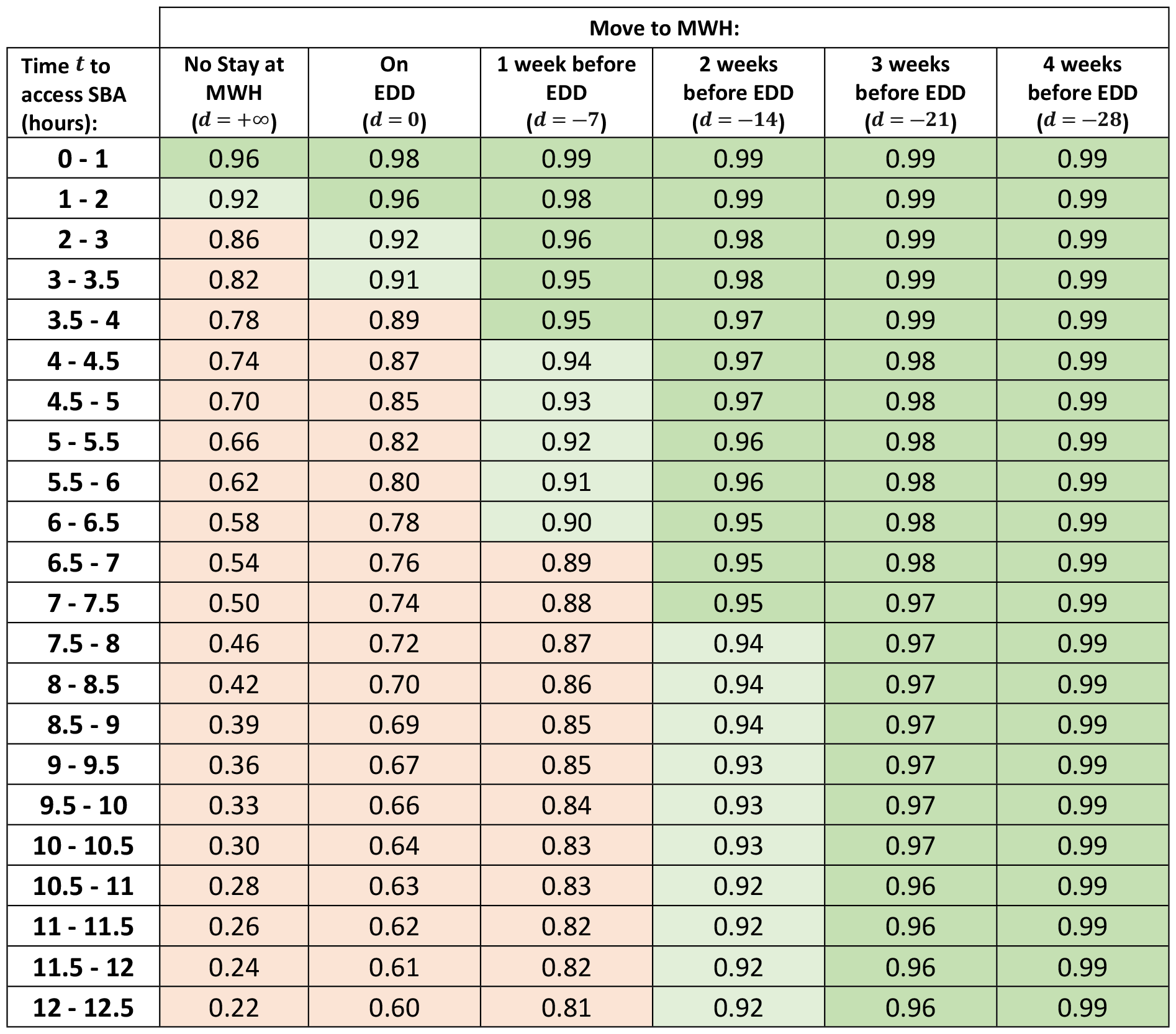
Estimated probability of skilled birth attendance ***P***(SBA) computed with mathematical model defined in §2 and input data defined in §3.1-2 (nulliparas). The regions highlighted with colours correspond to the cases ***P***(SBA) < 0.9 (salmon), 0.9 < ***P***(SBA) < 0.95 (light green) and P(SBA)> 0.95 (dark green).

| Time $t$ to<br>access SBA<br>(hours): | Move to MWH: | | | | | |
| --- | --- | --- | --- | --- | --- | --- |
| | No Stay at<br>MWH<br>($d = +\infty$) | On<br>EDD<br>($d = 0$) | 1 week before<br>EDD<br>($d = -7$) | 2 weeks<br>before EDD<br>($d = -14$) | 3 weeks<br>before EDD<br>($d = -21$) | 4 weeks<br>before EDD<br>($d = -28$) |
| 0 - 1 | 0.96 | 0.98 | 0.99 | 0.99 | 0.99 | 0.99 |
| 1 - 2 | 0.92 | 0.96 | 0.98 | 0.99 | 0.99 | 0.99 |
| 2 - 3 | 0.86 | 0.92 | 0.96 | 0.98 | 0.99 | 0.99 |
| 3 - 3.5 | 0.82 | 0.91 | 0.95 | 0.98 | 0.99 | 0.99 |
| 3.5 - 4 | 0.78 | 0.89 | 0.95 | 0.97 | 0.99 | 0.99 |
| 4 - 4.5 | 0.74 | 0.87 | 0.94 | 0.97 | 0.98 | 0.99 |
| 4.5 - 5 | 0.70 | 0.85 | 0.93 | 0.97 | 0.98 | 0.99 |
| 5 - 5.5 | 0.66 | 0.82 | 0.92 | 0.96 | 0.98 | 0.99 |
| 5.5 - 6 | 0.62 | 0.80 | 0.91 | 0.96 | 0.98 | 0.99 |
| 6 - 6.5 | 0.58 | 0.78 | 0.90 | 0.95 | 0.98 | 0.99 |
| 6.5 - 7 | 0.54 | 0.76 | 0.89 | 0.95 | 0.98 | 0.99 |
| 7 - 7.5 | 0.50 | 0.74 | 0.88 | 0.95 | 0.97 | 0.99 |
| 7.5 - 8 | 0.46 | 0.72 | 0.87 | 0.94 | 0.97 | 0.99 |
| 8 - 8.5 | 0.42 | 0.70 | 0.86 | 0.94 | 0.97 | 0.99 |
| 8.5 - 9 | 0.39 | 0.69 | 0.85 | 0.94 | 0.97 | 0.99 |
| 9 - 9.5 | 0.36 | 0.67 | 0.85 | 0.93 | 0.97 | 0.99 |
| 9.5 - 10 | 0.33 | 0.66 | 0.84 | 0.93 | 0.97 | 0.99 |
| 10 - 10.5 | 0.30 | 0.64 | 0.83 | 0.93 | 0.97 | 0.99 |
| 10.5 - 11 | 0.28 | 0.63 | 0.83 | 0.92 | 0.96 | 0.99 |
| 11 - 11.5 | 0.26 | 0.62 | 0.82 | 0.92 | 0.96 | 0.99 |
| 11.5 - 12 | 0.24 | 0.61 | 0.82 | 0.92 | 0.96 | 0.99 |
| 12 - 12.5 | 0.22 | 0.60 | 0.81 | 0.92 | 0.96 | 0.99 |

Consistent with the interpretation of Equation (1) provided in §2, the coloured regions highlighted in Tables 1 and 2 show how the risk of delivering without an SBA for a woman with a given parity living in a specific location (i.e., on a given row of either Table 1 or 2) can be mitigated until it reaches a desired threshold by increasing the time before EDD when she moves to an MWH (i.e., by considering intervention scenarios captured by columns further to the right in Tables 1 and 2 until an acceptably high value of ***P***(SBA) is found).

**Table 2:** Estimated probability of skilled birth attendance ***P***(SBA) computed with mathematical model defined in §2 and input data defined in §3.1-2 (multiparas). The regions highlighted with colours correspond to the cases ***P***(SBA) < 0.9 (salmon), 0.9 < ***P***(SBA) < 0.95 (light green) and ***P***(SBA)> 0.95 (dark green).

| Time $t$ to access SBA (hours): | Move to MWH: | | | | | |
| --- | --- | --- | --- | --- | --- | --- |
| | No Stay at MWH ($d = +\infty$) | On EDD ($d = 0$) | 1 week before EDD ($d = -7$) | 2 weeks before EDD ($d = -14$) | 3 weeks before EDD ($d = -21$) | 4 weeks before EDD ($d = -28$) |
| 0 - 1 | 0.93 | 0.96 | 0.98 | 0.99 | 0.99 | 0.99 |
| 1 - 2 | 0.82 | 0.91 | 0.95 | 0.98 | 0.99 | 0.99 |
| 2 - 3 | 0.69 | 0.84 | 0.92 | 0.96 | 0.98 | 0.99 |
| 3 - 3.5 | 0.62 | 0.80 | 0.91 | 0.96 | 0.98 | 0.99 |
| 3.5 - 4 | 0.56 | 0.77 | 0.89 | 0.95 | 0.98 | 0.99 |
| 4 - 4.5 | 0.50 | 0.74 | 0.88 | 0.95 | 0.97 | 0.99 |
| 4.5 - 5 | 0.44 | 0.71 | 0.87 | 0.94 | 0.97 | 0.99 |
| 5 - 5.5 | 0.39 | 0.69 | 0.85 | 0.94 | 0.97 | 0.99 |
| 5.5 - 6 | 0.35 | 0.66 | 0.84 | 0.93 | 0.97 | 0.99 |
| 6 - 6.5 | 0.30 | 0.64 | 0.83 | 0.93 | 0.97 | 0.99 |
| 6.5 - 7 | 0.26 | 0.62 | 0.82 | 0.92 | 0.96 | 0.99 |
| 7 - 7.5 | 0.23 | 0.60 | 0.81 | 0.92 | 0.96 | 0.99 |
| 7.5 - 8 | 0.19 | 0.59 | 0.81 | 0.92 | 0.96 | 0.99 |
| 8 - 8.5 | 0.16 | 0.57 | 0.80 | 0.91 | 0.96 | 0.99 |
| 8.5 - 9 | 0.14 | 0.56 | 0.79 | 0.91 | 0.96 | 0.99 |
| 9 - 9.5 | 0.12 | 0.55 | 0.79 | 0.91 | 0.96 | 0.99 |
| 9.5 - 10 | 0.10 | 0.54 | 0.78 | 0.91 | 0.96 | 0.99 |
| 10 - 10.5 | 0.08 | 0.53 | 0.78 | 0.90 | 0.96 | 0.99 |
| 10.5 - 11 | 0.06 | 0.52 | 0.78 | 0.90 | 0.95 | 0.99 |
| 11 - 11.5 | 0.05 | 0.51 | 0.77 | 0.90 | 0.95 | 0.99 |
| 11.5 - 12 | 0.04 | 0.51 | 0.77 | 0.90 | 0.95 | 0.99 |
| 12 - 12.5 | 0.03 | 0.51 | 0.77 | 0.90 | 0.95 | 0.99 |

## 4. Physical Device Implementation

The physical device to be described now is designed to facilitate the availability and consideration of the key output information provided by the mathematical model described earlier as part of the antenatal visits hopefully preceding birth for women living in low resource environments. As such its main intended user is the care professional conducting the antenatal consultation. This device essentially embodies the data shown in Tables 1 and 2, with a physical design meant to facilitate the correct consultation and understanding of these data. Its design is inspired from the widespread “pregnancy wheel” device commonly used to predict EDD based on LMP, which some of our intended users will be familiar with.

This device is composed of two independent sides, specifically a blue side for nulliparas and a purple side for multiparas, each involving two components. These two components are concentric discs: a smaller one positioned on top referred to as the ‘upper wheel,’ and a larger one beneath referred to as the ‘lower wheel,’ with relevant information only printed on the front (user-facing) side of these two wheels. The centers of these two wheels are aligned and connected so that the upper wheel may freely rotate around this common center / axis while remaining connected to the lower wheel; this aspect of the physical design is identical to that of the pregnancy wheel used to predict EDD from the LMP date just mentioned. The backs of the two lower wheels of each side may be permanently attached to each other, so that the information corresponding to nulliparas and multiparas may be separately accessed from the two independent sides of a single device.

The lower wheel (Figures 3B and 3D) contains all the data embodied by the device, which is effectively identical to the data contained in Tables 1 (Figure 3B) and 2 (Figure 3D).

**Figure 3:**
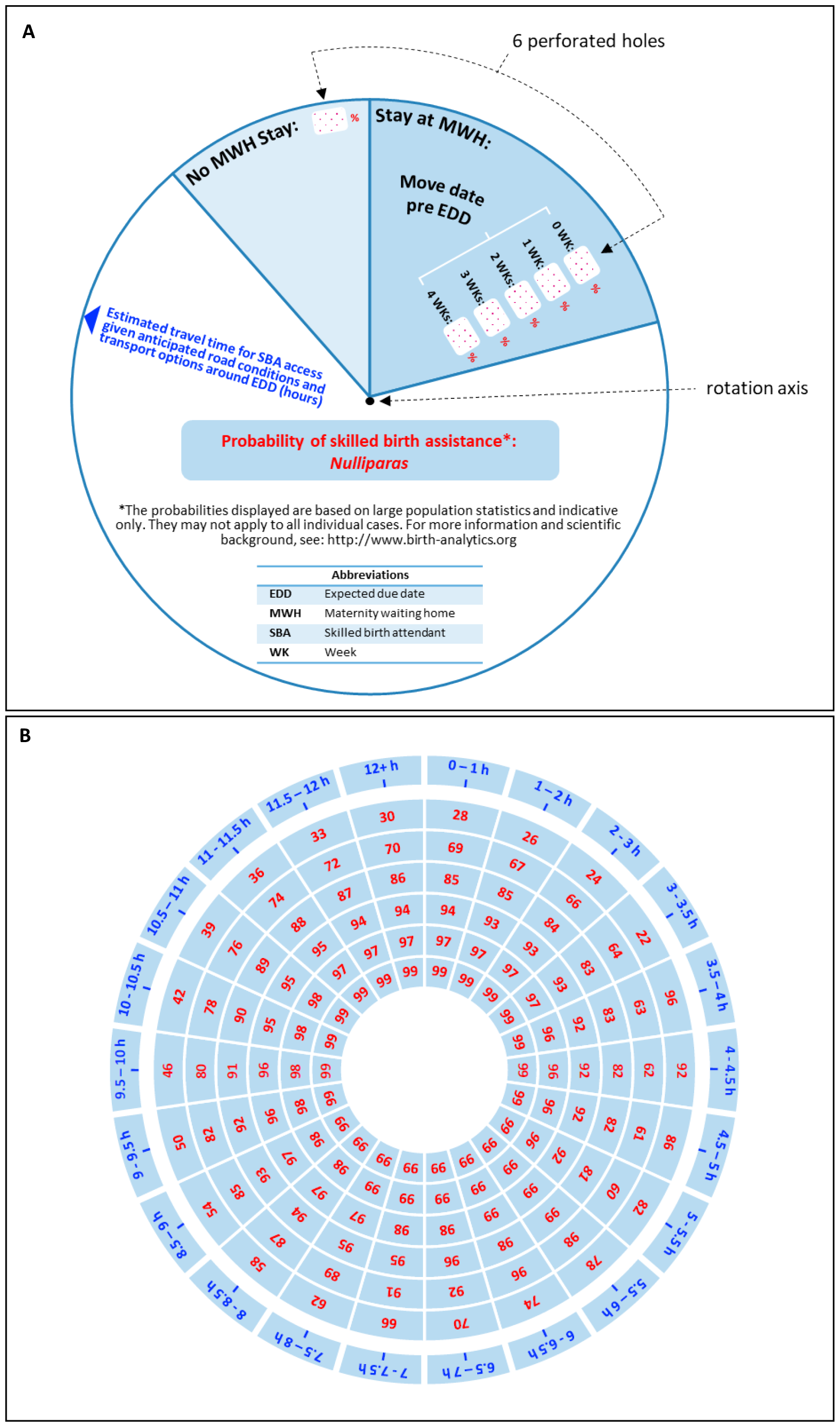
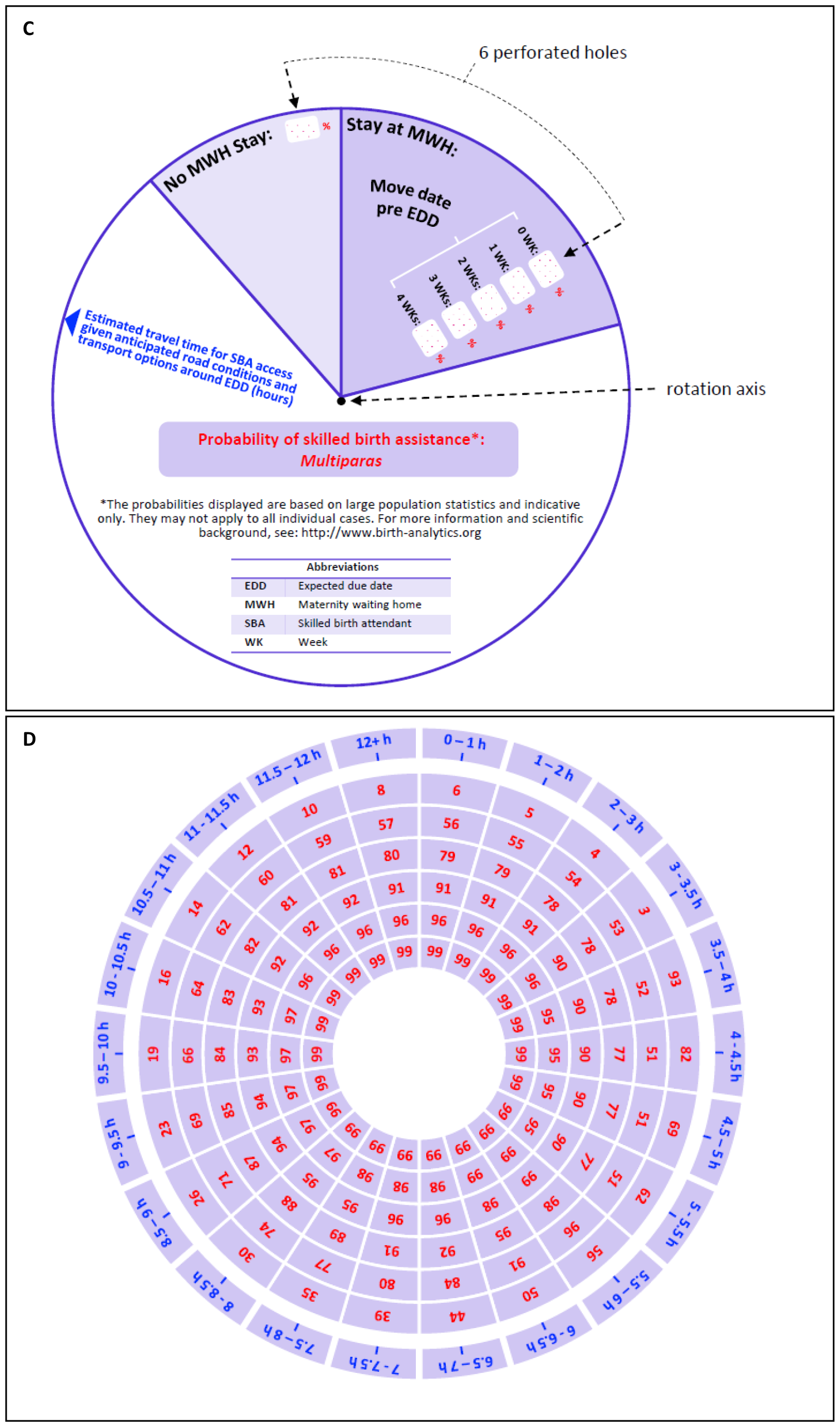
Graphical and geometric design of the four main components of the proposed physical device. Panel A: Upper wheel for nulliparas; Panel B: Lower wheel for nulliparas Panel C: Upper wheel for multiparas; Panel D: Lower wheel for multiparas

Specifically, the data contained in each of the 7 columns of Table 1 or 2 is displayed on the lower wheel alongside a concentric circle, with the data display circle radius decreasing as one considers the columns from left to right in the Table. For example, the outer circle of data contains the labels of access time categories in the first column of Tables 1 and 2, while the smallest / inner circle of data contains the predicted values of ***P***(SBA) for a move to an MWH 4 weeks before EDD in the last column of Table 1 and 2. Furthermore, each of these concentric circles of data is appropriately rotated around the wheel center in order to appropriately match the design of the upper wheel.

The upper wheel (Figures 3A and 3C) has a diameter smaller than that of the outer / largest circle of data on the lower wheel but larger than that of the second largest circle of data on the lower wheel. As a result the upper wheel conceals all of the data on the lower wheel except the access time interval label data on its outer edge and six particular ***P***(SBA) output data points which are visible and highlighted to the user through six appropriately placed and dimensioned holes / perforations on that wheel (these correspond to the 2^nd^ to 7^th^ columns of Tables 1 and 2). The upper wheel contains the definition of all the concepts and variables featured on the device, some disclaimer and reference information as well as an alignment arrow. This arrow allows / helps the user to rotate the upper wheel in relation to the lower wheel so that the ***P***(SBA) output data made visible through the perforated holes corresponds to the appropriate selected access time interval for the particular woman for whom the device is being used (see Figure 4 for illustration).

**Figure 4:**
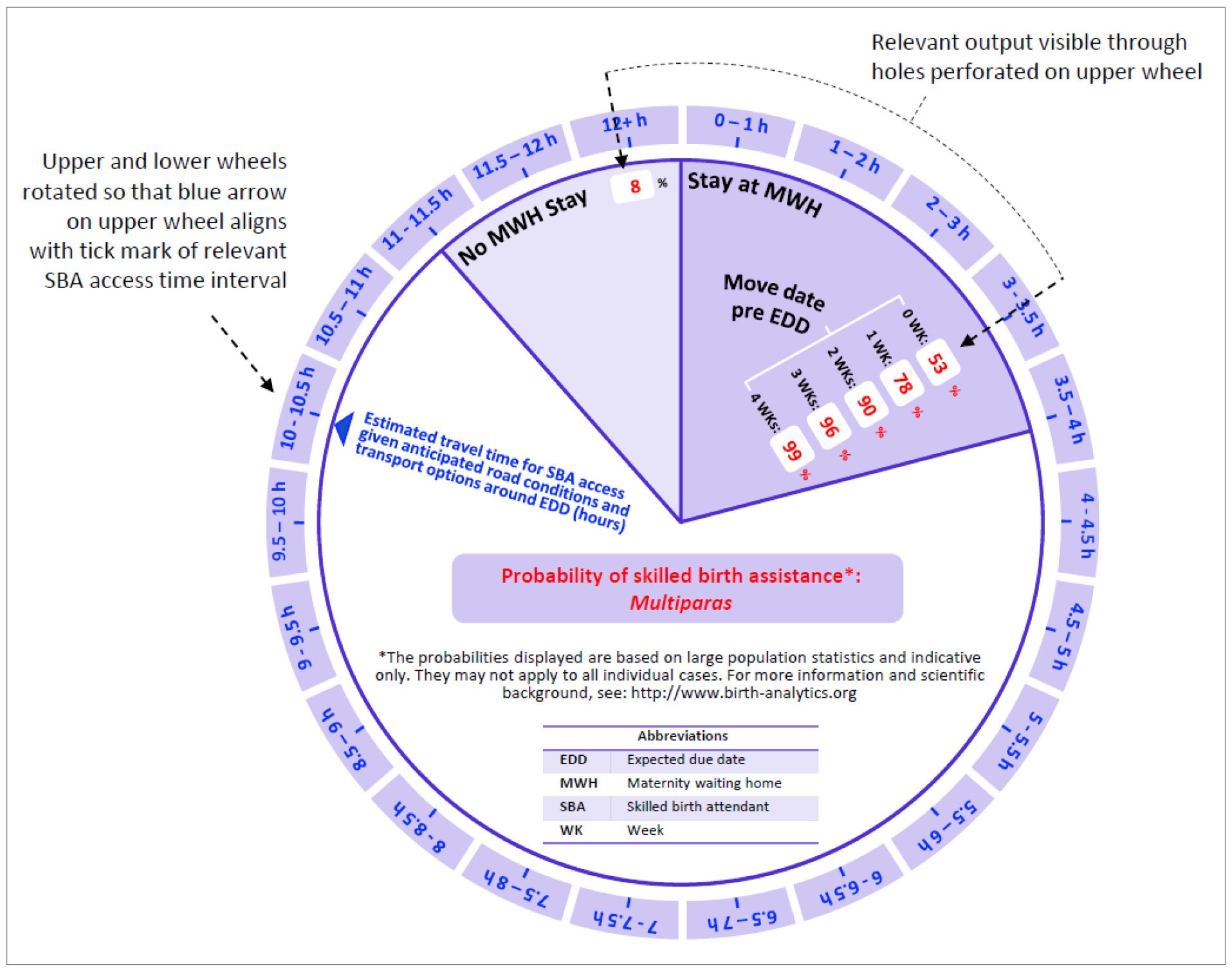
Example of the proposed physical device use for a multiparous woman living approximatively 10h15min away from the nearest skilled birth attendant given anticipated road conditions and transport availability around the time of her EDD. In this example the blue arrow of the upper wheel has been aligned with the middle tick mark of the SBA access time interval 10-10.5 hours on the outer edge of the lower wheel and the visible / highlighted ***P***(SBA) values correspond to those found in the second to seventh columns of Table 2 for the corresponding row, namely ***P***(SBA) = 8% without MWH stay and ***P***(SBA) = 53%, 78%, 90%, 96% or 99% with a MWH move date 0, 7, 14, 21 or 28 days before EDD, respectively.

## 5. Discussion

Equation (1) provides a systematic and quantitative individual prediction of the risk that a woman’s active phase of labour will begin without a skilled birth attendant based on some of her personal circumstances (parity, anticipated time to access SBA), and the extent to which that risk may be reduced by moving to a MWH at a specific time in advance of her EDD; these predictions are based on a simple and explicit mathematical model. As such this work provides a theoretical complement to the extensive existing literature reporting empirical assessments of the prevalence of SBA^3-4^, maternal and neonatal mortality by place of delivery^36^ and MWH use and benefits^13-16^.

The predictive accuracy of this model in the field is yet to be evaluated, and there are multiple reasons why it could be less than perfect. Among others, the cohorts of pregnant women used to estimate latent phase duration and labour onset distributions in the studies currently adopted as main data input to our model^8,9^ may have different characteristics than the population of women to whom this risk assessment model is applied in the field, and these differences may be material; our model may not capture all the relevant patient characteristics with a predictable impact on latent phase duration or labour onset distribution, in particular it explicitly captures no specific aspect of the clinical profile besides parity, and does not capture potential additional uncertainty around EDD arising when the LMP date is unknown. It also ignores the potential correlation between the date of labour onset and the latent phase duration and any uncertainty surrounding the predicted time to access an SBA upon labour onset. Finally, the definition of latent phase duration used in the studies providing that input of our model ignores the time that pregnant women, their families and/or care providers may take to make decisions relevant to obstetric care access upon the realisation that labour has started.

In the envisioned implementation setting for this model, predictive accuracy issues would arguably be most harmful in the case of substantial under-estimation of care access risk, potentially leading pregnant women or care providers to forego important / necessary care access arrangements. This consideration motivates our choice of the Friedman studies as the main source of our latent phase duration estimates, because these studies appear to report the shortest latent phase durations within the related available literature^8-12^.

Substantial over-estimation of predicted care access risk might primarily entail a loss of output credibility, which seems secondary to the potential health costs associated with under-estimation. The present modelling and input data choices reflect our attempt to strike a balance between these costs while recognizing the practical appeal of a simple model. We also stress that in the envisioned implementation setting, quantitative risk prediction errors would only be material if substantial enough to affect their qualitative interpretation.

Despite highlighted limitations, the care access risk assessment model and implementing physical device presented here may thus still be useful. To that end these tools should be used with appropriate perspective and sensitivity, as only one input in a broader interaction between pregnant woman and care providers that also includes a competent review of all relevant individual patient circumstances including medical profile and family situation. Indeed, this work is motivated by our field observations that individual care access plan considerations (as opposed to clinical aspects) are not systematically discussed as part of antenatal consultations in rural resource-limited settings, let alone supported by formal evidence-based guidance or risk assessment. In this context some practical implementation of our model adapted for use during an antenatal consultation, such as the physical device described in §4, may prompt more systematic and extensive discussions of perinatal care access arrangements and a heightened awareness of their importance by both care providers and pregnant women living in remote locations. Practical implementation of this work may also heighten the awareness of MWH stay as a possible intervention and/or of its benefits. It may also facilitate the design and consistent implementation of more specific and effective recommendations for MWH stay, which could be beneficial^37^. This may be achieved through simple rules whereby the earliest MWH move date resulting in a minimum specified ***P***(SBA) value is recommended (in the case illustrated by Figure 4 for example, a minimum specified value of ***P***(SBA) ≥ 90% in all cases would result in an individual recommendation of moving to the MWH 2 weeks before EDD). In areas where MWH bed capacity is restricted, one might adjust that minimum threshold value of ***P***(SBA) in order to limit the possibility of MWH overcrowding and associated negative consequences^38^. Although MWH stay is the only intervention explicitly considered by this model, the predicted risk of delivery without SBA provided for the scenario without MWH stay may also motivate the advice or planning of alternative types of care access arrangements (e.g., alternative temporary residence, transportation access arrangements, engagement of local care professional) by either care professionals or individual women and their families.

Future related work may seek to evaluate the accuracy of this mathematical risk assessment model; enhance this model by capturing additional relevant factor (EDD prediction accuracy, SBA access time uncertainty, further medical or other patient characteristics, etc.); develop formal antenatal consultation guidelines leveraging this model; evaluate the field impact of this tool and/or usage guidelines; and leverage this model for designing local MWH use recommendations and health facility network planning.

## Data Availability

All relevant data are within the manuscript and its Supporting Information files.

https://www.dropbox.com/scl/fi/lz9htuqkmlij29rx20vvu/Data-Tables.xlsx?rlkey=6e7tiy3n6l6mkvu9i6t06mq38&dl=0

## Abbreviations

The following table contains the abbreviations used in this document:

ADD: Actual Delivery Date
EDD: Expected Due Date
LMP: Last Menstrual Period
MWH: Maternity Waiting Home
SBA: Skilled Birth Attendant
SD: Standard Deviation

## Notes

### Competing Interest Statement

The authors have declared no competing interest.

### Funding Statement

Yes

